# Proposal of a vaccination needs index to prioritize municipal interventions in Brazil

**DOI:** 10.1101/2023.09.09.23295306

**Authors:** Fredi Alexander Diaz-Quijano, Isaac Negretto Schrarstzhaupt, Francieli Fontana Sutile Tardetti Fantinato, Lely Stella Guzmán-Barrera, Julio Croda

## Abstract

**Introduction:** To address the health needs of a population, it is essential to integrate several indicators that are expressed on non-comparable scales. Vaccination is a crucial aspect of public health, and prioritizing vaccination needs through immunization registries can help allocate resources and funding to integral approaches.

**Objective:** To propose and implement an index of vaccination needs (VNI) based on national administrative data on tracer vaccines for children, born between 2019 and 2022, to help prioritize Brazilian municipalities.

**Methodology:** We analyzed data from 5,570 municipalities in Brazil. The VNI integrated indicators of vaccination coverage and the number of susceptible children regarding tracer vaccines for children including DTP – Diphtheria-Tetanus-Pertussis (third dose), polio (third dose), and MMR – Measles-Mumps and Rubella (second dose). We assessed how the VNI could identify a group of municipalities in challenging situations to improve vaccination.

**Results:** Compared with criteria based only on the absolute number of unvaccinated children (susceptible children) or the vaccination coverage, the group prioritized for having high VNI exhibited more similar proportions of large, medium, and small municipalities. Additionally, this group included more municipalities located in the Legal Amazon region and Special Sanitary Indigenous Districts than groups prioritized with the other criteria. The VNI also outperformed the other criteria to prioritize municipalities that differed from the rest regarding the Gini coefficient.

**Conclusion:** The proposed VNI can facilitate the identification of populations that need differentiated interventions to prevent the resurgence of eliminated or controlled vaccine-preventable diseases.

## INTRODUCTION

To acknowledge health needs we often need to integrate several indicators that are expressed into non-comparable scales [1,2]. Some indexes are been used to represent health needs by aggregating standardized variables, usually after they are transformed to obtain distributions that, when compared, represent the variation of population according to domains of interest [2]. The resulting metrics can then be used to prioritize the funding and resource allocation to integral health approaches or to reorientate intervention strategies for population access to services [3].

Concerning the vaccination needs, the prioritization should take into account the coverage of the vaccines identified by PAHO-WHO as tracers for children under one year of age and one year of age according to the basic schedule worldwide. Besides considering several vaccines, another challenge is the nature of the indicator. We could use indicators such as the number of unvaccinated children (susceptible) or consider the vaccination coverage.

The absolute number of unvaccinated people directly represents the resource needs and affects the national indicators. However, the prioritization based on an absolute indicator would end up focusing on large capitals and systematically excluding smaller cities even if they have lower vaccine coverage [4]. On the other hand, the prioritization based exclusively on vaccination coverage would not consider the importance of the number of unvaccinated children and would undermine large groups of them that exist in big cities [5].

In this study, we proposed and applied a method to obtain an index of vaccination needs (VNI) considering three tracer vaccines of the routine schedule expected to be administrated in the first two years of age, integrating the absolute number of unvaccinated children and the correspondent vaccination coverage. To this end, we considered the complete primary schedule (3 doses) for Polio and Diphteria-Tetanus-Pertussis (DTP), and two doses for Measles-Mumps-Rubella (MMR) vaccine in children born between 2019 and 2022, using open data from Brazil’s National Immunization Program.

## METHODS

In this work, we proposed the construction of an index based on data recorded in the country’s official vaccination information system, following steps similar to those that have been used in the construction of other social science indexes [6,7]. The steps include the selection of the component indicators, the data gathering, the evaluation of the distributions, transformation, standardization, aggregation, and application to establish an order of the analysis units.

### Indicator selection

As available and conceptually relevant for public health, we considered tracers the primary schema including three doses for DTP and Polio vaccines, and two doses for MMR in children up to 4 years of age, in Brazil for the year 2022. Thus, we chose the following indicators:

- Number of unvaccinated children: In each city, we calculated the apparent number of children by age without the doses considered in the primary scheme.
- Vaccine coverage: In each city, we also calculated the proportion of children with the primary schema doses of each vaccine.

### Data source and indicator calculation

To calculate the vaccination coverage, the numerator and denominator were, respectively, the administered doses and the population for each age. Administered doses were obtained from the TABNET database, which is a platform with open health data. Filters to select the immunobiological corresponding to each vaccine were defined in consensus with the National Immunization Program (Supplementary material: Box S1). Population estimates were obtained from the National Institute of Geography and Statistics (IBGE) and the Brazilian Ministry of Health.

For each reference dose of the vaccines (*v*), the administered doses expected to accumulate in each cohort defined simple ages in each municipality were added. The ratio between the sum of those doses and the population estimates, for each age and municipality (*P*_*i,m*_: population of the age group “*i*” in the municipality “*m*”), was considered the specific coverage for each age-municipality (*C*_*v,i,m*_) and was truncated to 100%. The number of unvaccinated (susceptible) children specific to each year of age and municipality (*S*_*v,i,m*_) was calculated as *S*_*v,i,m*_ = (1 − *C*_*v,i,m*_) * *P*_*i,m*_

The coverage in the zero-to-four years’ group (*C*_*v,0-4,m*_) was calculated as the complement of the proportion of unvaccinated children of the corresponding age group. That proportion was calculated as the ratio between the sum of unvaccinated children for each age and the sum of the resident population for the same age. For example, the DTP vaccination coverage in the population of zero to four years in each municipality was calculated as:

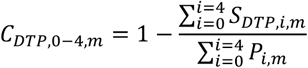

The absolute indicator would be the sum of unvaccinated children (*S*_*m,0-4,DTP*_), which would correspond to:

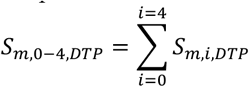

### Vaccination Needs Index (VNI) construction

The indicators of each vaccine were graphically analyzed to evaluate its distribution in the 5,570 Brazilian municipalities. Both the number of unvaccinated children and the vaccination coverage had asymmetrical distributions, for which transformations to minimize the difference of those distributions to a normal shape were explored. The number of unvaccinated children was transformed by adding a single unit and obtaining the natural logarithm of that result:

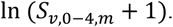

For the vaccination coverages, beyond the search for symmetry, the transformation included the inversion of the signal so that the sense could be the same as the indicator of susceptible children. The chosen transformation was:

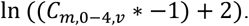

Each resulting variable was standardized by subtracting the average and dividing by its standard deviation. In this way, we obtained transformed and standardized measures (or Z-scores) for each absolute and relative indicator of each vaccine, all of them with a mean equal to zero and standard deviation equal to 1. As we summed the indicators of each vaccine, we obtained specific VNIs. The sum of the three vaccine-specific VNIs was considered as the consolidated VNI.

### Validation

In the absence of a gold standard to validate the VNI, we focused on assessing characteristics that we considered to be expected in a group of prioritized municipalities, such as diversity in population size or challenging situations to achieve high coverage. So, we described the characteristics of the municipalities that would be prioritized for belonging to the 5% (n=278) with the highest VNI. As alternative references, we also described the characteristics of municipalities chosen with two other criteria of prioritization:

1. The 278 cities with the highest susceptible (unvaccinated) average, considering the three vaccines studied;
2. The 278 cities with the lowest coverage average of the same vaccines.

Regarding the characteristics assessed, we expected that the prioritized group included both large and small cities, for having a high number of unvaccinated children or low vaccination coverages. The size of municipalities was classified according to population estimates as “large” (>100000 inhabitants), “medium” (between 25000 but lower than 100000 inhabitants), or “small” (<25000 inhabitants). Additionally, we expected that populations with more challenges to obtain high vaccination coverage, such as border municipalities, Legal Amazon cities, and Special Indigenous Sanitary Districts (DSEIs) have a good chance of being prioritized. Legal Amazon is a territory defined by Law 1,806, of 01/06/1953, which includes 772 municipalities in nine Brazilian states [8]. The DSEI is the decentralized structure that answers for the management of the Indigenous Health System in Brazil. There are currently 34 DSEIs defined according to the geographical occupation of indigenous communities, which does not follow the political State division [9]. Finally, considering it plausible that the cities with more vaccination needs are also the ones with greater socioeconomic inequality [10,11], we compared the Gini coefficient between prioritized and non-prioritized municipalities. The Gini coefficient

Both the distribution of unvaccinated children and the coverage of these vaccines were highly asymmetric across Brazilian municipalities (left and center columns of Figure 1). However, with the transformations, we obtained more symmetric vaccine-specific VNIs (Figure 1, right column). When consolidated into a single index, we observed an even more symmetric distribution, resembling a normal distribution (Figure 2). The values of this consolidated VNI ranged from -15.3 to +18.4, and in the 278 municipalities with the highest values the VNI was greater than 7.425.

**Figure 1.**
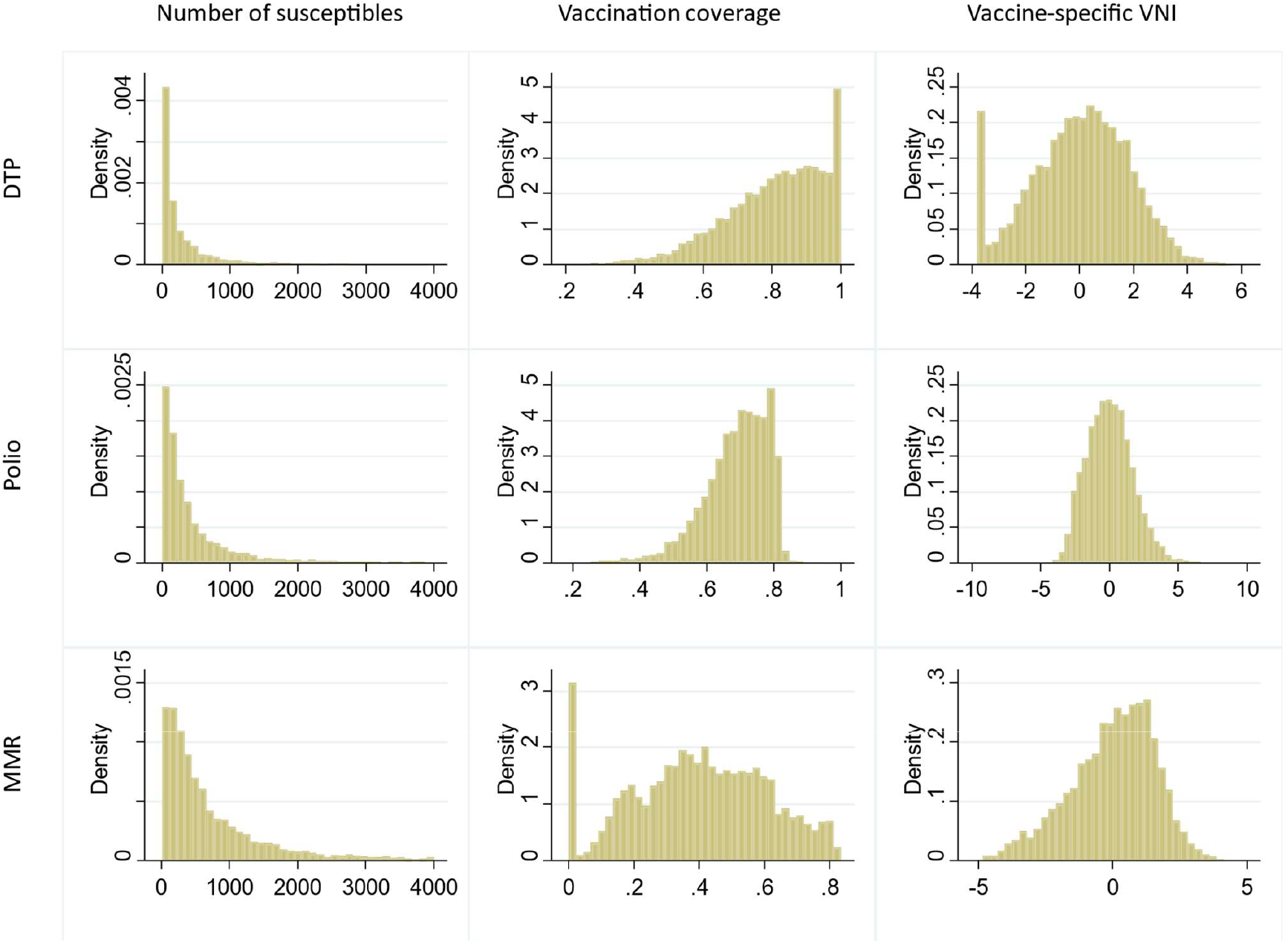
Distribution of number of unvaccinated children, vaccinations coverages and specific Vaccination Needs Index for Diphtheria-Tetanus-Pertussis, Polio and Measles-Mumps-Rubella in Brazil, 2022.

**Figure 2.**
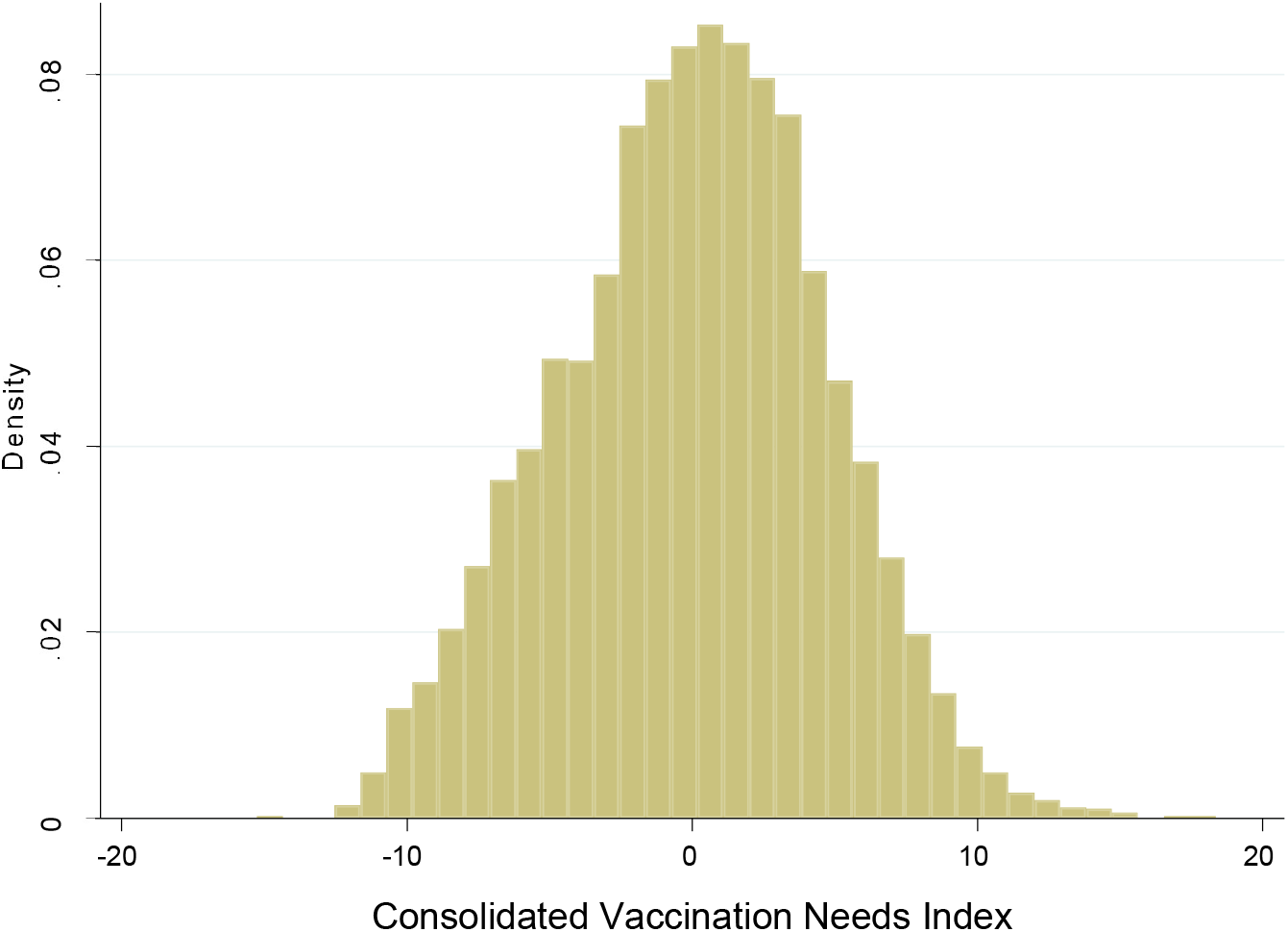
Distribution of a consolidated Vaccination Needs Index calculated for 5,570 municipalities in Brazil, 2022.

When comparing prioritization strategies, we observed that the 278 municipalities corresponding to the top 5% with the highest average number of measures inequality in the income distribution on a scale from zero to one [12,13]. A value of zero reflects perfect equality, where all income or assets are equal, while a Gini coefficient of one (or 100%) reflects the greatest inequality.

## RESULTS

For the year 2022, there were approximately 14.7 million children between zero and four years old (more than 2.9 million children in each single age category). In this population, we estimated coverage rates for the DTP (three doses), polio (three doses), and MMR (two doses) vaccines to be 79%, 67.7%, and 46.1%, respectively. Consequently, we calculated the number of unvaccinated children of approximately 3 million, 4.8 million, and 7.9 million for these vaccines, respectively. Although the number of unvaccinated children had medians that ranged from 132 to 490, this indicator reached into the hundreds of thousands (Table 1). Specifically, in São Paulo (SP) city, we calculated 149754 children without the third dose of DTP, 268849 without the third dose of polio, and 322336 without the second dose of MMR.

**Table 1:**
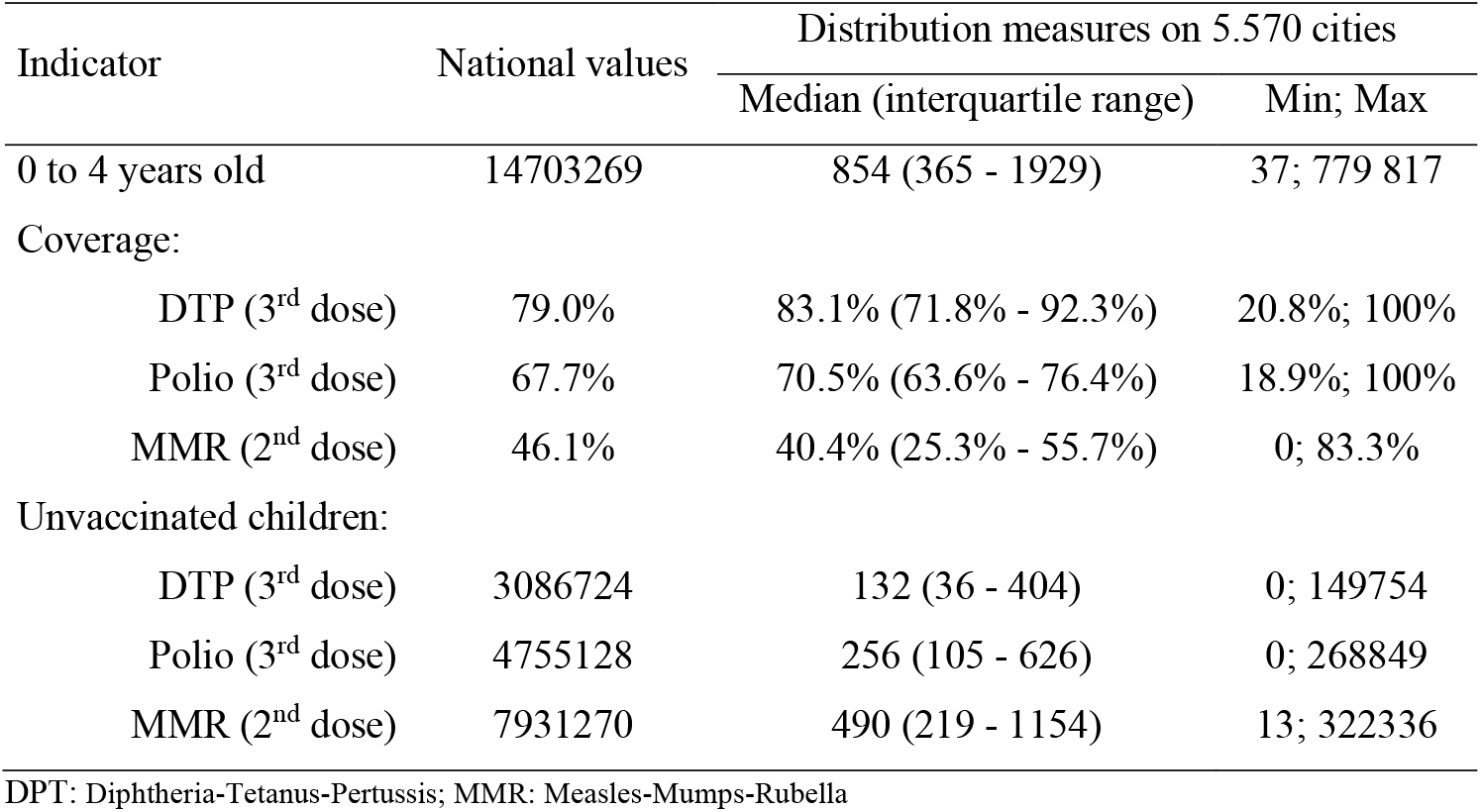
Vaccination coverage and unvaccinated children estimated for Diphtheria-Tetanus-Pertussis, Polio, and Measles-Mumps-Rubella vaccines, Brazil, 2022.

| Indicator | National values | Distribution measures on 5.570 cities |  |
| --- | --- | --- | --- |
|  |  | Median (interquartile range) | Min; Max |
| 0 to 4 years old | 14703269 | 854 (365 - 1929) | 37; 779 817 |
| Coverage: |  |  |  |
| DTP (3<sup>rd</sup> dose) | 79.0% | 83.1% (71.8% - 92.3%) | 20.8%; 100% |
| Polio (3<sup>rd</sup> dose) | 67.7% | 70.5% (63.6% - 76.4%) | 18.9%; 100% |
| MMR (2<sup>nd</sup> dose) | 46.1% | 40.4% (25.3% - 55.7%) | 0; 83.3% |
| Unvaccinated children: |  |  |  |
| DTP (3<sup>rd</sup> dose) | 3086724 | 132 (36 - 404) | 0; 149754 |
| Polio (3<sup>rd</sup> dose) | 4755128 | 256 (105 - 626) | 0; 268849 |
| MMR (2<sup>nd</sup> dose) | 7931270 | 490 (219 - 1154) | 13; 322336 |

DPT: Diphtheria-Tetanus-Pertussis; MMR: Measles-Mumps-Rubella

unvaccinated children were primarily large and did not include small ones. On the other hand, the 278 municipalities with the lowest coverage were mainly small (76.3%). Conversely, the group with the highest VNI had more similar proportions of large, medium, and small municipalities. Additionally, the group with the highest VNI had a higher frequency of municipalities located in the Legal Amazon region and Special Sanitary Indigenous Districts. The number of border cities was similar between the groups chosen based on coverage and VNI (21 and 20, respectively), but higher than the group chosen based on the number of unvaccinated children (Table 2).

**Table 2:**
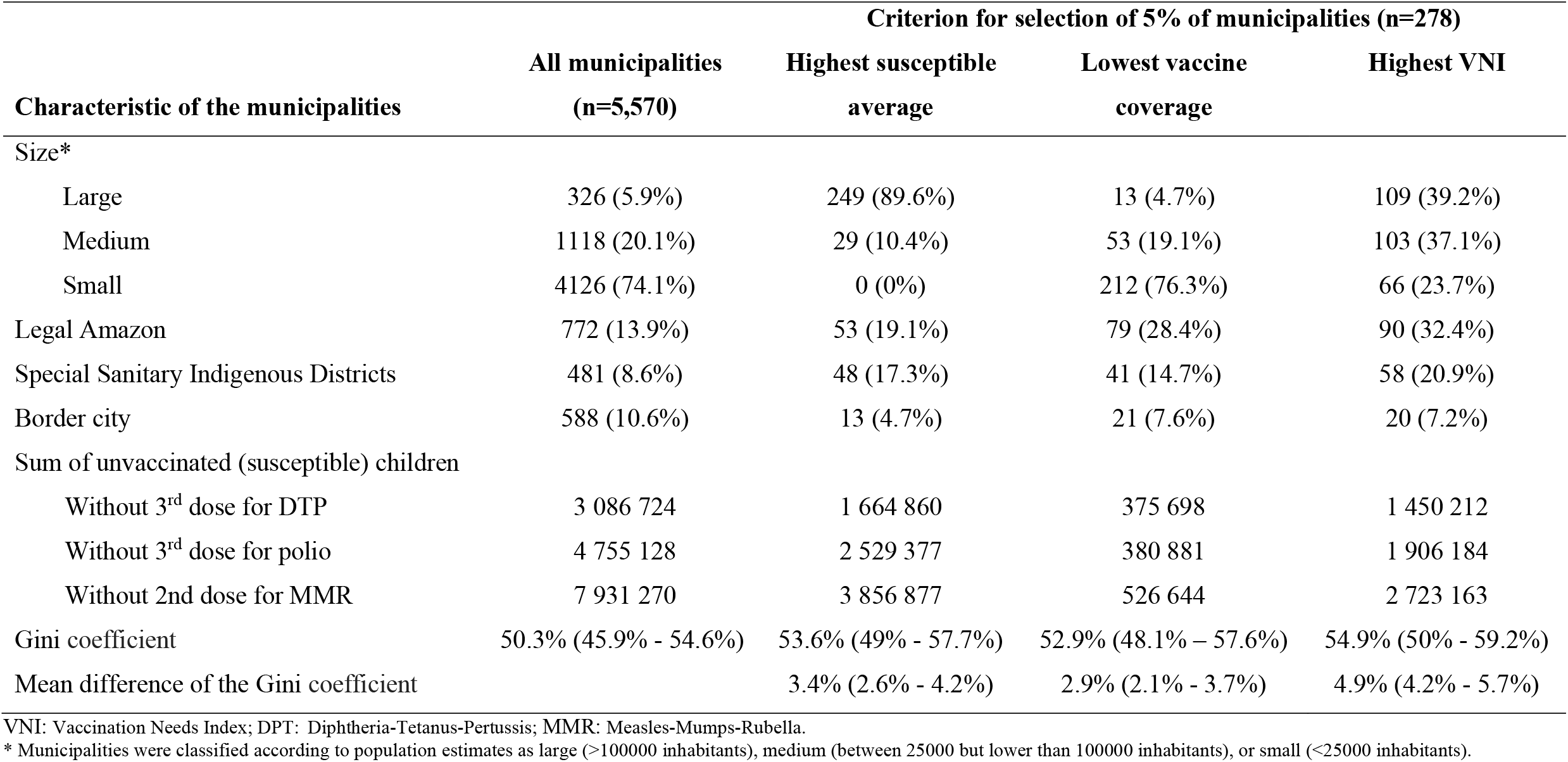
Validation of methods to prioritize populations according to vaccine needs.

The number of children without the reference doses was higher in municipalities chosen based on the number of unvaccinated children (as expected). However, municipalities with the highest VNI had relatively close values to this latter group. On the other hand, municipalities prioritized due to its low coverage had a much smaller number of unvaccinated children. Finally, regarding the inequality, municipalities chosen based on VNI had a higher average Gini coefficient differing by 4.9% from the rest of the municipalities. Although the other two prioritization criteria also led to differences in the Gini coefficient, these differences were smaller than observed with the VNI criterion (Table 2).

## DISCUSSION

Prioritization is essential for the efficient use of public health resources. However, choosing a metric to prioritize populations regarding vaccination needs is a methodological challenge. Absolut indicators such as the number of unvaccinated individuals are directly related to the resource requirements, including the number of doses and healthcare personnel, among others. On the other hand, the relative measure of coverage is important to avoid the exclusion of remote communities. Additionally, to achieve herd or collective immunity, the number of immune individuals should be distributed evenly [14,15], so achieving and maintaining high coverage in all municipalities can prevent the formation of susceptible clusters where outbreaks can begin. In addition, in smaller municipalities with low coverage, interventions with fewer resources can have a relatively greater impact by reducing the risk of vaccine-preventable diseases.

The proposed VNI in this study integrated absolute and relative indicators of essential vaccines. In the validation, this index allowed for increased representation within the prioritized group of municipalities with characteristics that suggest vulnerability, such as those with indigenous districts or regions in border areas, also encompassing a high number of unvaccinated people. Moreover, the VNI outperformed other evaluated prioritization strategies in identifying a group of municipalities that differed from the rest in terms of the Gini coefficient.

We consider that the limitations of the proposed VNI are linked to those of the indicators used for its construction, as well as the data recorded in the national vaccination information system. For example, the number of doses may not necessarily represent the number of immunized individuals. On the other hand, without an individualized registry of people, it is not possible to perform corrections like those for mortality in vaccinated and unvaccinated individuals. However, we expect the classification errors to be non-differential and therefore not affect the final prioritization order.

While other metrics can be developed to prioritize, including other social, economic, and other health aspects, we believe that this index has several advantages. For instance, it is very specific to the vaccination situation at the municipal level and can be easily constructed using systematically collected information. In addition, VNI can be used in a variety of ways. For example, several categories can be established for intervention levels, as in the case of the 200 municipalities with the highest VNI representing approximately one-third of the unvaccinated population with the reference doses (see “Supplemental Excel File”). On the other hand, by adding the following 1,000 cities, a total of 1,200 priority municipalities with the highest VNI include approximately two-thirds of the population without the reference doses for DPT, polio, and MMR in the country. In this way, decision-makers can apply different categorizations to plan activities with different levels of intervention and efficiently enhance vaccination access of the target population.

We believe that the proposed index is innovative. While indexes composed of indicators of diverse nature have been used in other scenarios, prioritization for improving vaccination coverage has so far considered relative or absolute values, but in isolation [16–18]. There are also mathematical models in the literature for vaccine prioritization, but none calculated a single index [19–21]. In this way, the VNI can be seen as a tool to integrate the needs of the person rather than to meet the goal of a specific vaccine. Moreover, we consider that the proposed methodology can be applied to other populations and adding other vaccines, depending on the requirements in the prioritization process.

In conclusion, the proposed INV integrated indicators of different natures that represent the vaccination situation according to the scheme indicated by age. This approach can facilitate the identification of populations that need differentiated interventions to improve their health situation and prevent the resurgence of eliminated or controlled diseases that are preventable with vaccines. This method can be adapted and adopted by any country that, based on its databases and system, wants to prioritize its target populations at the municipal management level.

## Data Availability

All data produced in the present work are contained in the manuscript.

## Authors’ contribution

FADQ participated in the conception and design of the study, devised the vaccination needs index, performed data analysis, and prepared the first version of the manuscript. INS made substantial contributions to data acquisition, FSTFF LG, and JC made significant conceptual insights for the methodology. All authors discussed the results of the first version, provided relevant information for writing, performed revisions, and read and approved the final manuscript. Therefore, each author participated sufficiently in the work to take public responsibility for appropriate portions of the content and, thus, agreed to be accountable for all aspects in ensuring that questions related to the accuracy or integrity of any part of the work are appropriately investigated and resolved.

## SUPPLEMENTARY MATERIALS

### Box S1.

**Selections in the applied doses website to build the initial database.**

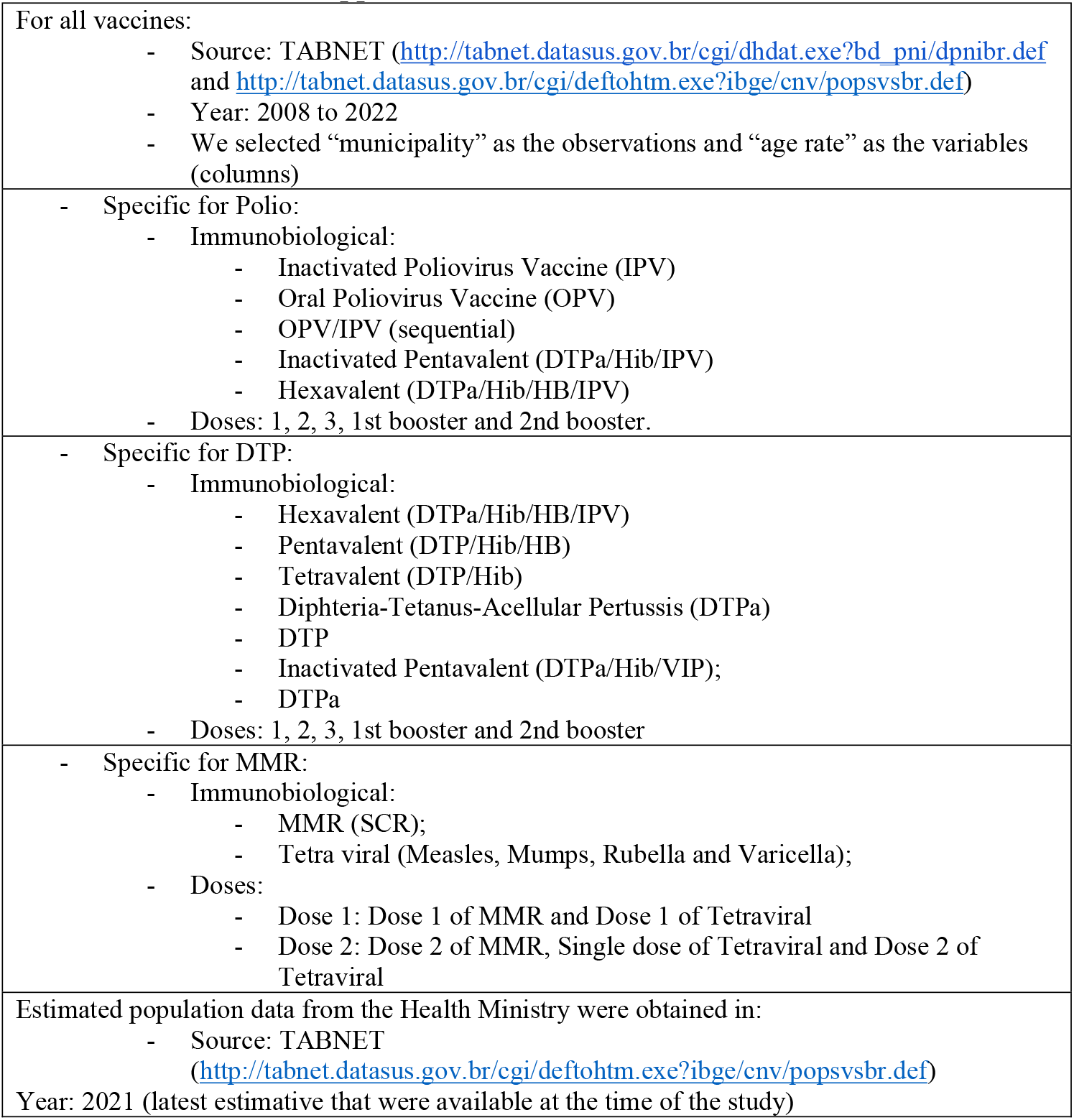

## REFERENCES

[1] Chen MK. Measuring need for health services: a proposed model. Med Care 1979;17:210–4. 10.1097/00005650-197902000-00011.

[2] Chen MK. A health needs index based on the health goals model. Public Health Rep 1983;98:181–4.

[3] Mooney A, Rives NWJ. Measures of community health status for health planning. Health Serv Res 1978;13:129–45.

[4] Murthy BP, Sterrett N, Weller D, Zell E, Reynolds L, Toblin RL, et al. Disparities in COVID-19 Vaccination Coverage Between Urban and Rural Counties - United States, December 14, 2020-April 10, 2021. MMWR Morb Mortal Wkly Rep 2021;70:759–64. 10.15585/mmwr.mm7020e3.

[5] Barbaro B, Brotherton JML. Assessing HPV vaccine coverage in Australia by geography and socioeconomic status : Aust N Z J Public Health 2014;38:419–23. 10.1111/1753-6405.12218.

[6] Zamankhani F, Abachizadeh K, Omidnia S, Abadi A, Hiedarnia MA. Composite social health index : Development and assessment in provinces of Iran Ministry of Health. Med J Islam Repub Iran 2019;33:1–6. 10.34171/mmjiri.33.78.

[7] Mousavi M, Nafei A, Rafei H, Shiani M, Mohammadi Gharehghani MA, Ghafuri R. Construction and Validation of Social Citizenship Index. Heal Scope 2021;10. 10.5812/jhealthscope.110283.

[8] IBGE A de N. IBGE atualiza Mapa da Amazônia Legal. 2020. https://agenciadenoticias.ibge.gov.br/agencia-sala-de-imprensa/2013-agencia-de-noticias/releases/28089-ibge-atualiza-mapa-da-amazonia-legal#:~:text=A Amazônia Legal foi instituída,a promoção de seu desenvolvimento.

[9] Ministério-da-Saúde. Distrito Sanitário Especial Indígena 2023. https://www.gov.br/saude/pt-br/composicao/sesai/estrutura/dsei.

[10] Tur-Sinai A, Gur-Arie R, Davidovitch N, Kopel E, Glazer Y, Anis E, et al. Vaccination uptake and income inequalities within a mass vaccination campaign. Isr J Health Policy Res 2019;8:1–8. 10.1186/s13584-019-0324-6.

[11] Zbiri S, Boukhalfa C. Inequality in COVID-19 vaccination in Africa. J Public Health Africa 2023;14:2353. 10.4081/jphia.2023.2353.

[12] De Maio FG. Income inequality measures. J Epidemiol Community Health 2007;61:849–52. 10.1136/jech.2006.052969.

[13] Asada Y, Hurley J, Norheim OF, Johri M. A three-stage approach to measuring health inequalities and inequities. Int J Equity Health 2014;13:1–13. 10.1186/s12939-014-0098-y.

[14] John TJ, Samuel R. Herd immunity and herd effect: new insights and definitions. Eur J Epidemiol 2000;16:601–6.

[15] Halloran ME. Concepts of transmission and dynamics. In: Thomas JC, Weber DJ, editors. Epidemiol. Methods Study Infect. Dis., New York: Oxford University Press; 2001, p. 56–85.

[16] Cunha NSP, Fahrat SCL, de Olinda RA, Braga ALF, Barbieri CLA, de Aguiar Pontes Pamplona Y, et al. Spatial analysis of vaccine coverage on the first year of life in the northeast of Brazil. BMC Public Health 2022;22:1204. 10.1186/s12889-022-13589-9.

[17] Akbarpour M, Budish E, Dworczak P, Kominers SD. An Economic Framework for Vaccine Prioritization*. Q J Econ 2023:qjad022. 10.1093/qje/qjad022.

[18] Pingali C, Yankey D, Elam-Evans LD, Markowitz LE, Williams CL, Fredua B, et al. National, Regional, State, and Selected Local Area Vaccination Coverage Among Adolescents Aged 13-17 Years - United States, 2020. MMWR Morb Mortal Wkly Rep 2021;70:1183–90. 10.15585/mmwr.mm7035a1.

[19] Chen S-I, Norman BA, Rajgopal J, Assi TM, Lee BY, Brown ST. A planning model for the WHO-EPI vaccine distribution network in developing countries. IIE Trans 2014;46:853–65. 10.1080/0740817X.2013.813094.

[20] Dastgoshade S, Shafiee M, Klibi W, Shishebori D. Social equity-based distribution networks design for the COVID-19 vaccine. Int J Prod Econ 2022;250:108684. 10.1016/j.ijpe.2022.108684.

[21] Lee EK, Li ZL, Liu YK, LeDuc J. Strategies for Vaccine Prioritization and Mass Dispensing. Vaccines 2021;9. 10.3390/vaccines9050506.

